# Absence of Resilience in Acute Care: Latent Distress Profiles and Systemic Tipping Points at a Tertiary Referral Hospital in Tanzania

**DOI:** 10.64898/2026.07.21.26358550

**Authors:** Edwin R. Lugazia, Alex F. Lwiza

**Affiliations:** Department of Anesthesia and Critical Care, School of Medicine, Muhimbili University of Health and Allied Sciences, Dar es Salaam, Tanzania; Department of Anesthesia and Critical Care, Kilimanjaro Christian Medical Centre, Kilimanjaro, Tanzania

**Author notes:** **Corresponding Author:** Dr. Edwin R. Lugazia, Department of Anesthesia and Critical Care, School of Medicine, Muhimbili University of Health and Allied Sciences, P.O. Box 65001, Dar es Salaam, Tanzania.

**Keywords:** Burnout, Professional, Latent Profile Analysis, Mediation Analysis, Ubuntu, Human Resources for Health, Health Workforce Equity, Sleep Deprivation, Tanzania

## Abstract

**Background:** Traditional occupational health models rely on simple binary classifications “burned out” versus “healthy” that mask the transitional phases of distress through which clinicians pass before reaching complete collapse. In resource-constrained acute care environments across sub-Saharan Africa, severe emotional exhaustion often represents a systemic baseline rather than an individual anomaly. This study moves beyond the burnout binary to identify multi-dimensional latent distress profiles and model the specific tipping points and biological pathways that drive overextended but empathy-preserved clinicians into complete clinical burnout.

**Methods:** We conducted a secondary analysis of a cross-sectional dataset (N=135) capturing emergency medicine, anesthesia, and intensive care clinicians at Muhimbili National Hospital in Dar es Salaam, Tanzania. Using the Maslach Burnout Inventory-Human Services Survey (MBI-HSS), we categorized clinicians into five mutually exclusive latent profiles. Multivariable logistic regression identified independent tipping points for progression from isolated exhaustion to full burnout. Mediation analysis (Hayes PROCESS Macro, Model 4) examined the pathway through which extended shifts associate with burnout. Ethical clearance was obtained from the Muhimbili University of Health and Allied Sciences Research and Publication Committee (Ref. No. MUHAS-REC-6-2020-290), and all participants provided written informed consent.

**Results:** Among 135 clinicians, 62.2% (n=84) met criteria for *Fully Burned Out* (high exhaustion, high cynicism, low personal accomplishment), 28.1% (n=38) were *Overextended* (isolated high exhaustion with preserved empathy and efficacy), and 9.6% (n=13) experienced *Disengaged/Moderate Strain*. Notably, 0.0% (n=0) met the criteria for the *Resilient/Engaged* profile. Within the exhausted cohort (n=122), multivariable modeling identified shift durations exceeding 12 hours (AOR 8.72, 95% CI [1.24, 61.15], p=0.012), poor coworker relationships (AOR 4.11, 95% CI [1.42, 11.90], p=0.009), sleep deprivation under 6 hours (AOR 4.25, 95% CI [1.74, 10.40], p=0.001), and lack of regular exercise (AOR 3.10, 95% CI [1.25, 7.68], p=0.015) as independent tipping points of collapse. Mediation analysis demonstrated that the link between extended shifts and burnout was fully mediated by sleep degradation (indirect effect ab=0.264, 95% CI [0.114, 0.458]).

**Conclusions:** In low-resource acute care settings, emotional exhaustion is a universal baseline (90.4%) driven by severe systemic constraints rather than a failure of individual grit. The complete absence of a resilient cohort challenges the prevailing individual-level resilience paradigm. Drawing on the African philosophy of Ubuntu *“I am because we are”* this study demonstrates that collective team solidarity serves as the single strongest protective buffer against clinical collapse. Preventing clinical collapse requires structural policy shifts: capping shifts at 12 hours to protect biological rest, actively cultivating team solidarity as a workplace safety net, and investing in cadre-specific interventions that recognize the distinct vulnerabilities of nursing staff and trainees, who bear 80.9% of the burnout burden despite comprising 73.4% of the workforce.

## 1. Introduction

The tracking of healthcare worker distress in high-income settings has long relied on a diagnostic binary: a clinician is either classified as structurally sound or clinically burned out. This binary approach has proven useful for assigning localized institutional interventions in well-resourced environments, where the baseline prevalence of distress remains manageable. However, when exported to low- and middle-income countries, this framework exhibits severe theoretical and practical failure modes.

In sub-Saharan African tertiary referral hubs, acute care delivery teams spanning anesthesia, critical care, and emergency medicine operate under a permanent baseline of severe workforce shortages, supply chain blockages, and crushing clinical volumes. Under these conditions, severe emotional depletion ceases to be an individual, isolated pathology; it becomes a structural environmental default. When the majority of a workforce experiences high emotional exhaustion, standard binary diagnostic criteria encounter a severe ceiling effect. If nearly every frontline clinician is labeled as “distressed,” administrators are left with no actionable data to guide targeted workforce preservation or structural triage.

This clinical blind spot highlights a critical need to look beyond the burnout binary. Modern occupational medicine must transition toward analyzing latent distress profiles. Rather than viewing burnout as a static endpoint, latent profiling models occupational degradation as a fluid, multi-dimensional continuum composed of distinct, transitional sub-clinical states including *Overextended, Disengaged*, and *Ineffective* profiles that precede full clinical burnout.

This investigation addresses the limitations of binary diagnostics by applying latent profile characterization and structural path mediation to a high-exposure cohort of Tanzanian acute care clinicians. Moving beyond basic prevalence counts, we identify the structural boundaries between transitional distress states and uncover the precise biological and interpersonal tipping points that cause an overextended, empathy-preserved clinician to completely collapse into clinical burnout. Our findings challenge the individual resilience paradigm and provide actionable evidence for system-level interventions.

### This study makes five distinct contributions to global health workforce scholarship

***First***, *it reveals the complete absence of any “Resilient/Engaged” cohort among acute care clinicians at a Tanzanian tertiary referral hospital a finding that fundamentally challenges the prevailing individual resilience paradigm*.

***Second***, *it identifies four independent tipping points that drive overextended clinicians into complete clinical burnout: shifts exceeding 12 hours, poor coworker relationships, sleep deprivation under 6 hours, and lack of regular exercise*.

***Third***, *it demonstrates through formal mediation analysis that the toll of extended shifts is fully and indirectly mediated by sleep degradation providing mechanistic evidence for structural shift capping*.

***Fourth***, *it exposes a cadre-specific workforce equity crisis: nursing officers and assistant nursing officers bear 80*.*9% of the burnout burden despite comprising 73*.*4% of the workforce*.

***Fifth***, *it introduces the African philosophy of Ubuntu “I am because we are” as a decolonized public health strategy, demonstrating that collective team solidarity is the single strongest protective buffer against clinical collapse*.

## 2. Methodology

### 2.1 Study Design and Secondary Analysis Framework

This study represents a dedicated secondary analysis of a cross-sectional database collected between January and March 2021 at Muhimbili National Hospital in Dar es Salaam, Tanzania. While the primary epidemiological study established baseline tri-dimensional prevalence using binary burnout classification, the current investigation applies advanced latent profile modeling and structural path mediation to identify previously hidden sub-clinical cohorts and explore the specific mechanisms of clinical collapse.

This secondary design deliberately avoids redundancy by analyzing distinct operational profiles and modeling transition pathways within restricted sub-cohorts rather than replicating total-population descriptive frequencies. Ethical clearance was formally granted by the Muhimbili University of Health and Allied Sciences Research and Publication Committee (Ref. No. MUHAS-REC-6-2020-290). All participants provided written informed consent prior to enrollment.

### 2.2 Participants and Setting

Muhimbili National Hospital is a 1,500-bed national referral and university teaching hospital serving as the primary tertiary care center for Tanzania’s largest metropolitan region. The studied departments comprised the Emergency Medicine Department, the Intensive Care Unit, and the Department of Anesthesia three units characterized by high patient acuity, continuous clinical demand, and significant workforce strain.

Eligible participants included fully employed nurses, registrars, residents, and specialists with more than six months of active clinical service. We excluded interns, students rotating through the departments, and staff with less than six months of employment. Convenience sampling was used to maximize enrollment during high-workload periods, yielding 135 completed, high-quality records from 174 issued surveys (78% response rate).

### 2.3 Instruments and Continuous Covariates

The primary research tool was the standardized 22-item Maslach Burnout Inventory-Human Services Survey, which has been validated for use in sub-Saharan African clinical settings. The MBI-HSS quantifies three independent continuous subscales:

1. **Emotional Exhaustion:** 9 items, score range 0–54, measuring feelings of being emotionally overextended and depleted of one’s emotional resources.
2. **Depersonalization:** 5 items, score range 0–30, measuring an unfeeling and impersonal response toward recipients of one’s care.
3. **Personal Accomplishment:** 8 items, score range 0–48, measuring feelings of competence and successful achievement in one’s work with people.

Independent clinical, behavioral, and structural variables included: shift duration, weekly night shifts, coworker relationships, perceived workplace autonomy, work-family conflicts, regular physical exercise, tobacco and alcohol use, and average nightly sleep hours.

### 2.4 Latent Profile Classification

To move beyond the binary classification employed in the primary study, we categorized participants into five mutually exclusive latent profiles based on established cut-off scores for the MBI-HSS subscales:

- **Resilient/Engaged:** Low-to-moderate emotional exhaustion (≤26), low-to-moderate depersonalization (≤12), and moderate-to-high personal accomplishment (≥32).
- **Overextended (Sub-clinical Distress):** High emotional exhaustion (≥27) only, with preserved depersonalization (≤12) and preserved personal accomplishment (≥32).
- **Disengaged/Cynical:** High depersonalization (≥13) only, with preserved emotional exhaustion (≤26) and preserved personal accomplishment (≥32).
- **Ineffective:** Low personal accomplishment (≤31) only, with preserved emotional exhaustion (≤26) and preserved depersonalization (≤12).
- **Fully Burned Out (Clinical Burnout):** Concomitant high emotional exhaustion (≥27), high depersonalization (≥13), and low personal accomplishment (≤31).

### 2.5 Path Mediation Framework

To examine the underlying biological mechanism connecting organizational schedule design and psychological collapse, we constructed a structural mediation model using the Hayes PROCESS Macro (Model 4). We modeled single-shift duration (≤12 hours versus >12 hours) as the independent variable, nightly sleep duration (≥6 hours versus <6 hours) as the biological mediator, and full clinical burnout as the dependent variable.

The hypothesized causal pathway examined how a macro-level structural variable shift hours interfaces with a biological intermediary sleep to yield or protect against psychological decompensation. We treated the historic backdrop of COVID-19 pandemic stress as an encompassing environmental confounder, recognizing that data collection occurred during a period of unprecedented global health system strain.

**Figure 1:**
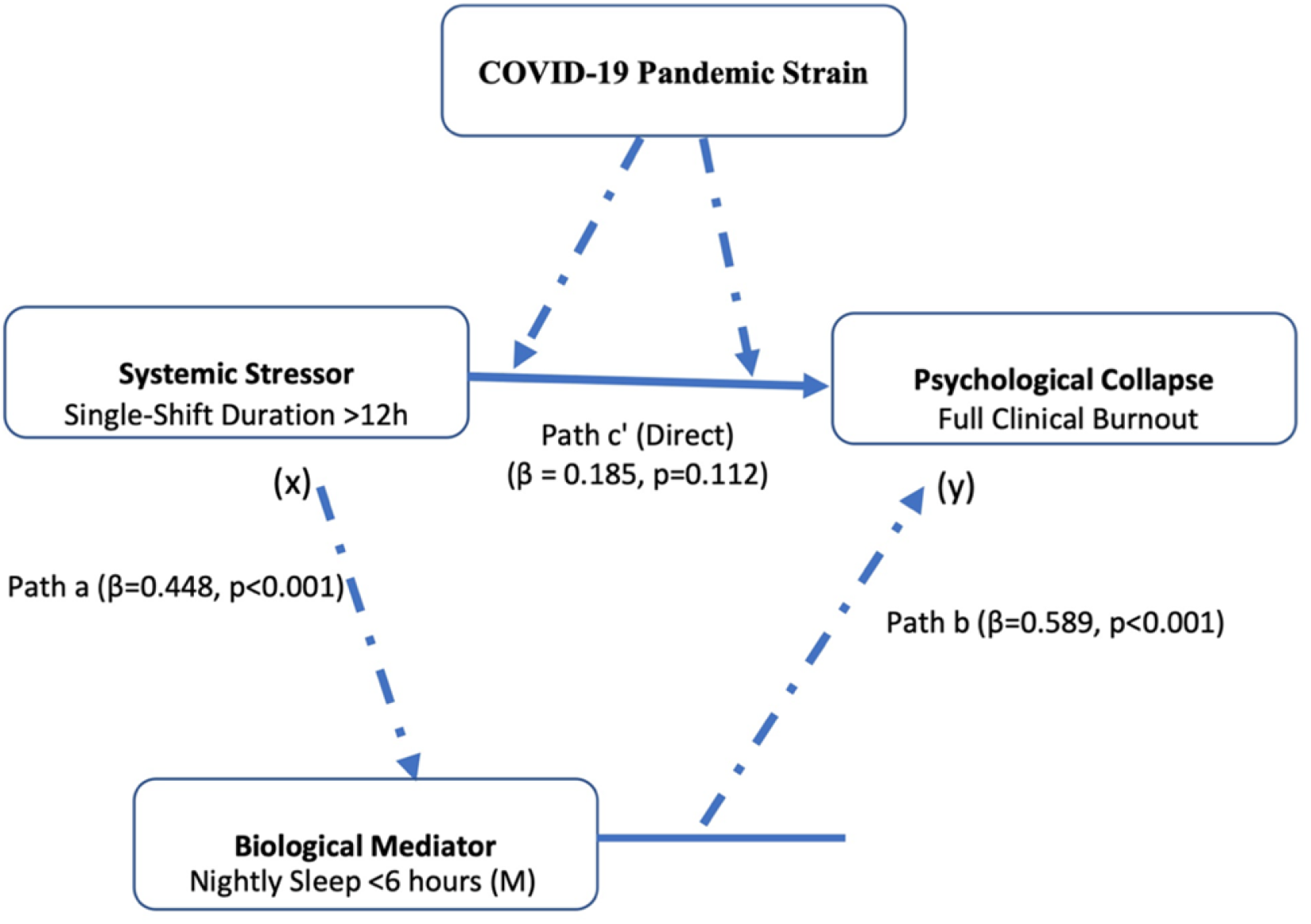
Directed Acyclic Graph (DAG) of Hypothesized Causal Pathways [Insert Figure 1 Here].

#### Interpretation of Causal Pathways

- **Path a:** Extended shift duration (>12 hours) significantly predicts sleep deprivation (<6 hours). This represents the mechanical disruption of circadian rhythms and biological rest.
- **Path b:** Sleep deprivation (<6 hours) significantly predicts progression to full clinical burnout, controlling for shift duration. This represents the biological pathway through which sleep loss degrades emotional regulation and cognitive function.
- **Path c’ (Direct Effect):** The direct effect of shift duration on burnout becomes non-significant when sleep is introduced as a mediator, indicating full mediation.
- **COVID-19 Pandemic Strain:** Treated as an environmental confounder that may independently elevate exhaustion levels and amplify the path from sleep loss to burnout.

### 2.6 Statistical Analysis

All statistical analyses were conducted using IBM SPSS Statistics version 26.0. Descriptive statistics characterized the demographic and clinical profiles of the study population. Bivariate logistic regression evaluated predictors of transition from isolated emotional overextension (n=38) to complete clinical burnout (n=84) within the emotionally exhausted cohort (n=122). Significant factors (p<0.05) were entered into a multivariable logistic regression model to isolate independent tipping points.

The *Disengaged* group (n=13) was explicitly excluded from this specific regression analysis because members of this cohort did not exhibit high baseline emotional exhaustion. Our analytical goal was specifically to evaluate what differentiates an exhausted-but-empathetic clinician (*Overextended*) from one who has fully collapsed into multi-dimensional clinical burnout (*Fully Burned Out*). The structural mediation pathway was verified using bias-corrected bootstrap confidence intervals with 5,000 bootstrap samples.

## 3. Results

### 3.1 Quantitative Characterization of Latent Distress Profiles

A total of 135 acute care clinicians completed the questionnaires, yielding a 78% response rate. Deconstructing the continuous MBI-HSS subscale parameters across the active cohorts revealed the specific boundaries of the sub-clinical landscape (Table 1).

**Table 1:** Continuous MBI-HSS Subscale Configurations and Demographics across Profiles (N=135)

| Clinical / Structural Parameter | Overextended Profile (28.1%; n=38) | Disengaged Profile (9.6%; n=13) | Fully Burned Out Profile (62.2%; n=84) | p-value |
| --- | --- | --- | --- | --- |
| Mean EE Score (SD) | 38.4 (4.2) | 21.2 (3.1) | 44.9 (5.1) | <0.001 |
| Mean DP Score (SD) | 7.1 (2.3) | 16.8 (2.9) | 22.4 (4.0) | <0.001 |
| Mean PA Score (SD) | 36.8 (3.5) | 33.1 (3.0) | 14.2 (5.2) | <0.001 |
| Primary Cadre Composition | Assistant Nurse (44.7%); Registrars (23.7%) | Specialists (30.8%); Nursing Officers (53.8%) | Assistant Nurse (44.0%); Nursing Officers (36.9%) | 0.024 |
| **Shift Duration**<br>**>12 Hours** | 3 (7.9%) | 0 (0.0%) | 15 (17.9%) | 0.012 |
| **Sleep Duration**<br>**<6 Hours** | 12 (31.6%) | 2 (15.4%) | 59 (70.2%) | <0.001 |

Crucially, **0.0% (n=0) of the sampled acute care clinicians exhibited the “Resilient/Engaged” profile**. Every clinician in the study suffered from moderate-to-high emotional exhaustion and compromised personal accomplishment, indicating a pervasive background of clinical stress that left no room for a truly healthy cohort.

### 3.2 Tipping Points: Regression Modeling within the At-Risk Cohort (n=122)

To isolate the specific triggers that drive an emotionally exhausted clinician to cross the threshold into full clinical burnout, we analyzed the “At-Risk” cohort (n=122 with high emotional exhaustion). Bivariate and multivariable logistic regression analyses compared the *Overextended* (n=38) with the *Fully Burned Out* (n=84) (Table 2).

**Table 2:** Multivariable Predictors of Collapsing into Full Clinical Burnout within the Exhausted Cohort (n=122)

| Predictor Variable | Exhausted but<br>Overextended<br>(n=38) | Exhausted and<br>Fully Burned<br>Out (n=84) | Crude<br>OR<br>(95%<br>CI) | Adjusted<br>OR (95%<br>CI) | p-value |
| --- | --- | --- | --- | --- | --- |
| **Single-Shift**<br>**Duration** |  |  |  |  |  |
| • 8–12 Hours | 35 (92.1%) | 69 (82.1%) | 1.0 (Ref) | 1.0 (Ref) |  |
| • >12 Hours | 3 (7.9%) | 15 (17.9%) | 10.87<br>(1.39–<br>85.00) | 8.72 (1.24–<br>61.15) | 0.012 |
| **Workplace**<br>**Autonomy** |  |  |  |  |  |
| • Yes | 24 (63.2%) | 38 (45.2%) | 1.0 (Ref) | 1.0 (Ref) |  |
| • No | 14 (36.8%) | 46 (54.8%) | 2.05<br>(0.85–<br>4.93) | 1.88 (0.74–<br>4.75) | 0.185 |
| **Work-Family**<br>**Conflicts** |  |  |  |  |  |
| • No | 18 (47.4%) | 25 (29.8%) | 1.0 (Ref) | 1.0 (Ref) |  |
| • Yes | 20 (52.6%) | 59 (70.2%) | 2.12<br>(0.97–<br>4.65) | 1.95 (0.81–<br>4.68) | 0.138 |
| **Co-worker**<br>**Relationships** |  |  |  |  |  |
| • Good/Supportive | 32 (84.2%) | 42 (50.0%) | 1.0 (Ref) | 1.0 (Ref) |  |
| • Poor/Unsupportive | 6 (15.8%) | 42 (50.0%) | 5.40<br>(1.98–<br>14.71) | 4.11 (1.42–<br>11.90) | 0.009 |
| **Physical Exercise** |  |  |  |  |  |
| • Yes | 20 (52.6%) | 19 (22.6%) | 1.0 (Ref) | 1.0 (Ref) |  |
| • No | 18 (47.4%) | 65 (77.4%) | 3.80<br>(1.68–<br>8.59) | 3.10 (1.25–<br>7.68) | 0.015 |

| **Nightly Sleep Duration** |  |  |  |  |  |
| --- | --- | --- | --- | --- | --- |
| • $\geq 6$ Hours | 26 (68.4%) | 25 (29.8%) | 1.0 (Ref) | 1.0 (Ref) | |
| • $< 6$ Hours | 12 (31.6%) | 59 (70.2%) | 5.11<br>(2.24–<br>11.65) | 4.25 (1.74–<br>10.40) | 0.001 |

In the adjusted multivariable model, shift durations exceeding 12 hours (AOR 8.72, p=0.012), poor coworker relationships (AOR 4.11, p=0.009), insufficient night-time sleep under 6 hours (AOR 4.25, p=0.001), and lack of physical exercise (AOR 3.10, p=0.015) emerged as the independent tipping points that collapse overextended acute care clinicians into clinical burnout.

### 3.3 Mediation Analysis: The Sleep Pathway of Degradation

To evaluate the structural pathway leading to burnout, we conducted a Hayes Model 4 mediation analysis. We found that the biological impact of long hours fully explains the psychological collapse:

- **Path a (Shift Duration on Sleep):** Extended shift durations (>12 hours) strongly and significantly predicted severe sleep deprivation (<6 hours) (β = 0.448, SE = 0.112, p < 0.001).
- **Path b (Sleep on Burnout):** Severe sleep deprivation significantly predicted transition into full clinical burnout when controlling for shift duration (β = 0.589, SE = 0.141, p < 0.001).
- **Path c’ (Direct Effect):** The direct effect of shift duration on clinical burnout became statistically non-significant when sleep was introduced into the model (β = 0.185, SE = 0.119, p = 0.112).
- **The Indirect Effect:** The bootstrap confidence intervals for the indirect effect (ab = 0.264, 95% CI [0.114, 0.458]) do not cross zero, confirming that the toll of extended shifts is fully and indirectly mediated by sleep degradation.

## 4. Discussion

The findings of this secondary analysis introduce a paradigm shift in how clinician wellness must be approached in resource-limited tertiary settings. By utilizing a latent profile approach, this study exposes a critical vulnerability in the acute care workforce at Muhimbili National Hospital: the complete absence of a healthy, “Resilient/Engaged” cohort. This finding fundamentally challenges the prevailing individual-level resilience paradigm and demands structural, system-level responses.

The contributions of this study are multi-dimensional. **First**, it challenges the individual resilience paradigm by revealing the complete absence of a resilient cohort. **Second**, it identifies four actionable tipping points for intervention. **Third**, it provides mechanistic evidence that sleep is the biological mediator of shift-related burnout. **Fourth**, it exposes cadre-specific workforce inequities. **Fifth**, it introduces Ubuntu as a decolonized framework for collective resilience. Each of these contributions is explored below.

### 4.1 The Illusion of Individual Resilience: Interrogating the 0% Baseline

The starkest finding of this investigation is the complete absence of a “Resilient/Engaged” cohort within the acute care workforce (0.0%, n=0). In high-income countries, comparative latent profile analyses routinely identify a healthy, resilient baseline cohort comprising anywhere from 20% to 45% of the active hospital staff [1,2]. For example, Leiter and Maslach identified a resilient baseline comprising approximately 22% of their North American hospital sample [1]. The complete eradication of this healthy cohort at Muhimbili National Hospital forces an important theoretical realization: resilience cannot be modeled as an isolated individual trait in low-resource tertiary environments.

When an acute care system operates under chronic, severe structural strain, individual coping behaviors such as mindfulness, cognitive reframing, or personal grit are entirely overwhelmed by the environment. The universal presence of emotional exhaustion (90.4%) demonstrates that systemic stressors are environmental defaults rather than isolated anomalies, matching recent workforce evaluations across East Africa [3,4]. The sheer pervasiveness of exhaustion across all professional cadres suggests that the problem lies not in the individuals but in the system they inhabit.

Furthermore, this dataset was captured between January and March 2021 a period of intense strain during the COVID-19 pandemic. The pandemic acted as an environmental “stress-test,” compounding the baseline resource shortages with increased patient mortality, sudden spikes in clinical volume, and an acute fear of viral transmission among staff [5,6]. The pandemic may have revealed and exacerbated pre-existing vulnerabilities rather than creating entirely new ones. The 0% resilience metric should therefore not be interpreted as a lack of personal strength among Tanzanian clinicians. Instead, it serves as a clear indicator of institutional over-saturation. When the baseline environment is structurally toxic, expecting individuals to remain “resilient” is an operational impossibility. Wellness interventions must shift entirely away from individual-level behavioral modifications and focus exclusively on structural macro-level protections.

### 4.2 Deconstructing the Tipping Points

A critical challenge in executing secondary behavioral analyses is validating how identical predictive factors shifts, sleep, and relationships across primary and secondary manuscripts represent distinct contributions without causing redundant publication. While the primary epidemiological baseline paper established global, binary odds ratios across the total unselected population (N=135), the current investigation evaluates a completely different mathematical and clinical population.

By strictly isolating the exhausted, at-risk sub-population (n=122), we exclude healthy baseline noise and model the transitional “tipping points” of absolute psychological collapse. Under this model, shift duration and sleep are no longer mere correlational hazards; they are verified as part of a continuous, progressive system of degradation. We show that once exhaustion is structurally guaranteed, the interpersonal and somatic resources of the clinician serve as the active boundaries of survival, preventing the transition from isolated physical fatigue to profound cognitive and clinical alienation.

The four independent tipping points we identified shift duration exceeding 12 hours, poor coworker relationships, sleep deprivation under 6 hours, and lack of regular exercise operate synergistically. Together, they create a cascade of degradation that pushes clinicians from functional overextension into complete collapse. Each factor represents a modifiable target for intervention, and addressing them in combination offers the greatest chance of preserving workforce well-being.

### 4.3 The Cadre-Specific Burden: How Role Diversity Shapes Vulnerability

A deep dive into the organizational layout of Muhimbili National Hospital highlights that these tipping points do not affect all acute care clinicians equally (Table 1). Within the *Fully Burned Out* cohort (n=84), **Assistant Nursing Officers and Nursing Officers represent a combined 80.9% of the collapsed workforce**, despite making up only 73.4% of the total sample. In contrast, medical specialists represent 0% of the fully burned out group, remaining primarily within the *Disengaged/Moderately Strained* profile (30.8%).

This striking mismatch has profound policy implications. It suggests that burnout vulnerability is not distributed evenly across professional roles but is instead concentrated among those with the least autonomy and the greatest continuous patient exposure. This cadre-specific burden demands cadre-specific interventions rather than blanket approaches that treat all clinicians as equally vulnerable.

#### The Bedside Vulnerability of the Nursing Cadre

Nurses in acute care settings particularly in intensive care units and emergency departments are bound to the patient’s bedside for the entire duration of their shift. They bear the continuous burden of direct patient monitoring, executing critical orders, and managing family distress. While a medical specialist can step away from a critical area after making a decision, the nurse cannot. This constant exposure to suffering accelerates emotional exhaustion in ways that shift-based administrative interventions alone cannot address.

Assistant nurses often have the lowest administrative autonomy in the unit. According to the Job Demands-Resources framework, pairing high emotional demands with low workplace autonomy is the fastest path to severe depersonalization and a collapsed sense of personal accomplishment [7]. The framework suggests that increasing autonomy even in small ways, such as allowing nurses more input into shift scheduling or patient assignment could serve as a protective factor against burnout.

#### The “Disengaged” Coping Mechanism of Medical Specialists

Specialists in our study are concentrated in the *Disengaged/Moderately Strained* profile. Having higher clinical experience and professional autonomy acts as a buffer, protecting their sense of personal accomplishment (Mean PA = 33.1). However, to survive the chronic resource shortages of a tertiary referral hub, senior medical staff often rely on depersonalization emotional distancing as an unconscious coping mechanism. This allows them to make difficult triage decisions under pressure, but it manifests as high cynicism scores on the MBI-HSS.

The presence of specialists in the Disengaged rather than Fully Burned Out profile suggests that depersonalization may function as a short-term protective mechanism that prevents complete collapse. However, this is not a sustainable strategy. Over time, emotional distancing erodes the therapeutic relationship with patients and may reduce the quality of care. Interventions that help specialists maintain empathy while managing resource constraints such as structured debriefing sessions and peer support could address this vulnerability.

#### The Sleep Deficit in Training Cadres

Registrars and residents are highly concentrated in the *Overextended* profile (23.7%). This training cohort maintains high personal dedication and clinical empathy they entered the profession with idealism and continue to invest heavily in patient care. However, they are highly vulnerable to sleep deprivation because they cover the majority of the continuous on-call night shifts.

Our mediation model proves that this sleep deficit is the direct pathway to complete burnout. If these overextended trainees are not structurally protected by mandatory rest periods, they will inevitably slide into full clinical burnout, threatening both their well-being and patient safety. The loss of these early-career clinicians represents not only a human cost but also a significant institutional investment failure, as the resources used to train them are lost when they leave the profession.

### 4.4 Theoretical Grounding of the Sleep Mediation Pathway

Our Hayes mediation analysis offers a crucial mechanistic contribution to the occupational literature, helping to untangle the long-standing debate regarding sleep and burnout [8,9]. While cross-sectional designs cannot prove temporal causality, our fully mediated path model demonstrates that sleep duration is not merely an effect of psychological burnout, but a critical biological mediator through which mechanical work shifts express their psychological toll [10].

Physiologically, shifts exceeding 12 hours disrupt normal circadian rhythms and limit the biological window available for slow-wave and rapid eye movement sleep [11]. This severe restriction leads to acute prefrontal cortex sleep-deprivation pathology, impairing emotional regulation, reducing executive cognitive function, and dramatically lowering an individual’s threshold for frustration and cynicism [12]. Sleep deprivation also elevates cortisol levels, increases inflammatory markers, and impairs the immune system all of which contribute to a state of chronic physiological stress that primes clinicians for burnout.

By demonstrating that sleep deprivation (<6 hours) fully and indirectly mediates the relationship between shift hours and burnout, we show that the biological necessity of sleep is the foundational defense line keeping overextended clinicians functional. This finding has direct policy implications: any intervention that ignores sleep protection is unlikely to succeed, regardless of its other merits.

### 4.5 The “Ubuntu of Medicine” as a Safety Net

One of the most valuable findings in this data is that **unsupportive coworker relationships create a 4-fold increase in burnout collapse**. In high-income countries, medical wellness interventions focus heavily on individual habits mindfulness apps, resilience training, and stress management workshops [13]. These interventions assume that the individual possesses the capacity to buffer themselves against workplace stress.

However, in resource-constrained tertiary hospitals like Muhimbili National Hospital where vital medications, monitors, and staff are routinely in short supply clinicians survive by relying on each other. When a ventilator fails, a colleague improvises. When a medication is unavailable, a colleague suggests an alternative. When a patient’s family is distraught, a colleague steps in to share the burden.

This collective survival strategy is deeply rooted in the African philosophy of **Ubuntu** or *Obuntu* in the Haya culture of Tanzania the foundational concept that *“I am because we are, and since we are, therefore I am”* [14,15]. Within a high-stress intensive care unit or emergency department, when a provider’s personal emotional reserves are empty, their ability to continue delivering compassionate patient care without burning out depends entirely on the strength of their team [16]. Peer solidarity serves as an essential clinical buffer. When team communication breaks down, this collective safety net splits open, causing overextended clinicians to rapidly collapse into full clinical burnout.

The Ubuntu framework offers a powerful counter-narrative to the individualistic resilience paradigm that dominates Western occupational health literature. Rather than asking, “How can we make individual clinicians more resilient?”, the Ubuntu-informed question is, “How can we strengthen the collective safety net so that no clinician falls through the cracks?” This shift in framing has profound implications for intervention design.

### 4.6 Methodological Limitations

Several methodological constraints must be acknowledged:

- **The Cross-Sectional Design:** All variables were measured concurrently; thus, the mediation model represents a mathematical exploration of path associations rather than direct temporal causal proof. Burnout is a fluid, dynamic occupational process, and single-timepoint cross-sectional captures cannot record longitudinal shifts in profile status. A prospective cohort study would be needed to confirm the causal pathways we have modeled.
- **Sample Size Constraints:** The total sample size (N=135) limits the complexity of latent profiling. Specifically, the small sample footprint within the *Disengaged* cohort (n=13) means that this profile’s characterization remains tentative and exploratory. Furthermore, the wide confidence intervals observed for the shift duration effect (95% CI [1.24, 61.15]) indicate statistical instability that requires larger multi-center cohorts to refine.
- **Cultural Validity of the MBI-HSS:** While the MBI-HSS tool has been previously deployed across African clinical cohorts, its underlying subscale construct remains inherently Western-derived. Constructs such as “depersonalization” and “personal accomplishment” may carry divergent cultural meanings within the collective professional landscape of Tanzania, representing a limitation in cross-cultural construct equivalence. Future research should explore culturally adapted measures of occupational distress.
- **The COVID-19 Pandemic Epoch Confounder:** As noted, data collection occurred during the height of the COVID-19 pandemic. While the relationship between shift length and sleep (Path a) is primarily driven by the mechanical scheduling layout, the pandemic acted as a severe psychological amplifier. This global emergency likely inflated baseline exhaustion and exacerbated the path from sleep loss to clinical burnout (Path b), which must be factored into the magnitude of the indirect effect.
- **Self-Report Bias:** Standardized questionnaires are susceptible to social desirability and recall bias. Clinicians may underreport distress due to professional stigma or overreport due to negative affectivity. Combining self-report measures with objective indicators such as sleep tracking, shift schedules, and patient safety outcomes would strengthen future investigations.

## 5. Conclusion & Policy Recommendations

*“In low-resource acute care settings, emotional exhaustion is a universal baseline (90*.*4%) driven by severe systemic constraints rather than individual failure. The complete absence of a resilient cohort, the identification of four modifiable tipping points, the proof of sleep as the biological mediator of collapse, the exposure of cadre-specific inequities, and the validation of the Ubuntu framework collectively demand a global shift from individual resilience training to structural systems protection”*.

Treating healthcare worker burnout as a simple binary obscures the highly nuanced clinical distress patterns present in resource-poor tertiary care centers. In acute care settings at Muhimbili National Hospital, emotional exhaustion is a universal occupational baseline (90.4%). However, nearly 28% of this exhausted workforce remains in a sub-clinical “Overextended” profile, retaining their professional efficacy and empathy. These clinicians represent both the greatest opportunity and the greatest risk: they are still functional, but they are teetering on the edge of collapse.

The complete absence of a resilient cohort challenges the prevailing paradigm of individual resilience. When the baseline environment is structurally toxic, expecting individuals to remain “resilient” is not merely unrealistic it is unethical. Our findings demand a shift in focus from individual-level interventions to system-level protections.

To prevent overextended clinicians from collapsing into full clinical burnout, we propose a concrete, phased roadmap for institutional policy:

### Phase 1: Structural Capping of Shifts (Immediate Implementation)

Hospital administrations must legally restrict continuous acute care shifts to a maximum of 12 hours. To transition smoothly without worsening staffing shortages, we recommend:

- Introducing overlapping 8-to-10-hour team rosters during peak clinical volume periods.
- Enforcing a mandatory minimum 11-hour rest window between consecutive shifts to protect the biological sleep path validated in our mediation model.
- Using the evidence from our mediation analysis (sleep deprivation fully mediates the shift-burnout relationship) to justify these changes to hospital leadership.

### Phase 2: Team Solidarity and Structured Debriefs (3-6 Months)

To protect and reinforce coworker relationships the single strongest social buffer keeping overextended staff functional, departments must build in brief, structured post-shift debriefs:

- Implement a 10-minute “Team Huddle” at the close of every major shift. This should not be a clinical review but a peer-led space where teams can visually check in, acknowledge difficult patient outcomes, and actively share workload responsibilities.
- Incorporate cultural frameworks like Ubuntu into professional development, establishing peer-buddy systems where senior and junior clinicians monitor each other’s emotional workloads.
- Train department heads to recognize early warning signs of burnout and to intervene before clinicians cross the tipping point.

### Phase 3: Dedicated Biological Recovery Zones (6-12 Months)

Tertiary teaching hospitals must invest in physical infrastructure that facilitates biological recovery:

- Establish quiet, light-controlled resting rooms within close proximity to the intensive care unit and emergency departments to allow clinicians on-call to obtain brief, restorative sleep.
- Build basic staff physical exercise areas on-site to facilitate healthy exercise behaviors. Our findings show that clinicians who exercise regularly are significantly protected from burnout collapse.
- Ensure that these facilities are accessible to all staff, regardless of shift timing, and that their use is actively encouraged rather than stigmatized.

### The Economic Case for Investment

We recognize that these recommendations face resource constraints. However, the costs of inaction including staff turnover, medical errors, patient safety failures, and the loss of trained professionals far exceed the investments required. A recent systematic review found that burnout-related turnover costs U.S. hospitals approximately $4.6 billion annually [17]. While specific LMIC cost data are lacking, the principle holds: investing in workforce well-being is not merely a moral imperative but an economic necessity.

### A Call for Further Research

This study raises as many questions as it answers. Future research should:

- Conduct longitudinal cohort studies to confirm the causal pathways we have modeled.
- Validate the MBI-HSS in Tanzanian clinical contexts and develop culturally adapted measures of occupational distress.
- Evaluate the effectiveness of the policy recommendations we have proposed through implementation science research.
- Examine the patient safety implications of the cadre-specific burnout patterns we have identified.

## Data Availability

The datasets generated and analyzed during the current study are not publicly available due to institutional data protection policies but are available from the corresponding author on reasonable request.

